# Effect of household boiling on ciprofloxacin and enrofloxacin residues in cow milk from Dhaka, Bangladesh: paired HPLC-UV screening and dietary exposure assessment

**DOI:** 10.64898/2026.08.18.26360522

**Authors:** Munir Ahmed, Md. Abdullahil Asif, Farhana Rinky, Md. Golam Mostafa, Mohammad Nazrul Islam Bhuiyan, Sadia Afrin, Taihan Ahmed, Asma Rahman

## Abstract

Boiling raw milk before drinking is a common practice in Bangladesh, but its impact on residue levels of ciprofloxacin and enrofloxacin is not well known. This study looked at these antibiotics in 120 raw cow milk samples from 12 dairy areas near Dhaka, collected between November 2023 and March 2024. Each sample was split into two parts: one tested as raw milk and the other after boiling at home for 15 minutes. Residues were checked using matrix-matched HPLC-UV.

Ciprofloxacin and enrofloxacin were found in 113 out of 120 samples (94.2%), and at least one of them was present in 117 samples (97.5%). We considered a level of 12.5 micrograms per liter or higher as detected. The average combined amount of these antibiotics dropped from 194.61 micrograms per kilogram in raw milk to 179.57 micrograms per kilogram after boiling, a decrease that was statistically significant (p < 0.001). However, the percentage of samples exceeding the EU maximum residue limit of 100 micrograms per kilogram only went down from 92.5% in raw milk to 90.0% after boiling, which was not statistically significant (p = 0.250).

Using national milk consumption data as an estimate of intake, the hazard index for adults was 0.360 for raw milk and 0.332 for boiled. Calculated with a 10 kg body weight, these values were 2.159 and 1.992, respectively. Overall, boiling at home did reduce the levels of ciprofloxacin and enrofloxacin, but it usually did not bring high-residue samples below the EU limit.

## 1. Introduction

Milk is a vital part of life in Bangladesh, providing essential nutrients such as protein, fat, vitamins, and calcium [1]. With population growth, urbanization, and changing eating habits, the demand for milk has risen [2,3]. Dairy farms and milk collection points near big cities like Dhaka have also expanded, supporting both nutrition and people’s livelihoods [2,3]. But with this growth, food safety becomes even more important. At every stage, from treating the animals and milking to transporting and selling the milk, chemicals and microbes can be picked up [4–6].

Antibiotics are commonly used to treat infections like mastitis and respiratory illnesses in dairy cows [5,7–11]. When used properly, they can be very helpful. But if doses are not carefully controlled, records are incomplete, veterinary supervision is missing, or milk is sold before the withdrawal period ends, residues could remain in the milk [5–7,10,12,13]. Using drugs responsibly and keeping accurate farm records helps prevent these residues [6,10,13].

The presence of antibiotic residues in milk is a concern for both consumers and the dairy industry [6,14,15]. These residues can cause allergic reactions, disrupt gut bacteria, have toxic effects, and contribute to antimicrobial resistance [6,14,16–18]. They can also interfere with the fermentation process used to make yogurt, cheese, and other dairy products [6,15]. Additionally, antibiotics from farm waste and manure can enter the environment, contributing to the increase of antimicrobial resistance [16,17,19].

Enrofloxacin (ENR) and ciprofloxacin (CIP) are antibiotics in the fluoroquinolone group that are important to watch for in veterinary residues [18,20–22]. ENR is given to animals and can turn into CIP, which remains active in the body [18,22]. Studies show that levels of both drugs usually go down as the withdrawal period increases [23]. Finding ENR and CIP in milk may mean that treatment or withdrawal steps weren’t always followed properly [6,10,13,23].

Regulatory limits and food safety guidelines answer different questions. In the European Union, the maximum residue limit (MRL) for cow’s milk is 100 µg/kg for the total of ENR and CIP [24]. We only use this as a reference point; it’s not a legal limit in Bangladesh. The World Health Organization’s JECFA set an acceptable daily intake of 0-2 µg/kg of body weight per day for enrofloxacin but didn’t set a specific MRL [25]. So, MRLs show how well residue controls are working, while exposure estimates help gauge how much people might ingest [24–26].

Milk is a tricky sample to analyze because it contains proteins, fats, salts, and other stuff that can affect testing [27–31]. Different methods have been used, like microbiological tests, HPLC, LC-MS/MS, and high-resolution mass spectrometry [20,27–36]. HPLC-UV is good for screening and measuring, but it’s less selective than LC-MS/MS [32,33,36]. That’s why in this study, we call the results screening estimates rather than confirmed identifications.

Reports of antibiotic residues in milk from Bangladesh and other countries vary a lot [12,32,33,35–46]. For example, Wang and colleagues found antibiotics in 10.6% of milk samples in Shanghai using high-resolution mass spectrometry [35]. Differences in results come from country, drug type, sample kind, and testing methods [32,33,35,36,38,44]. Because of this, results from one study shouldn’t be directly applied to another population.

Boiling is another important issue because many households boil raw milk before drinking it. Heating improves microbiological safety, but it does not always remove veterinary drug residues [1,47–52]. Quinolones can remain relatively stable during common heat treatments [47,51,52]. At the same time, uncovered boiling causes water loss, which can alter the measured concentration [47,49,51,52]. If milk volume or mass is not measured before and after heating, the result should be reported as a net concentration change rather than a true degradation rate [47,49,51,52].

This study looked at 120 milk samples collected from 12 areas around Dhaka. We tested both raw and boiled milk for CIP and ENR levels using HPLC-UV. We compared the two types of milk, checked for exceedances of EU-MRL standards, looked at regional differences, and estimated how much people might be exposed to these drugs through their diet. Since the samples were from specific regions and not a nationwide survey, the results shouldn’t be applied to all milk in Bangladesh.

## 2. Materials and methods

### 2.1 Study design, and sample collection

This study was conducted from November 2023 to March 2026 in 12 dairy areas near Dhaka, Bangladesh. Milk samples were taken from active collection points in each area. From each location, 10 raw cow milk samples were collected, for a total of 120 samples. The number of samples was chosen to cover all areas evenly during the study period and based on lab capacity. Since we didn’t do a nationwide sample size calculation, these results are specific to the areas studied and shouldn’t be seen as representative of all of Bangladesh.

Each original sample was split into two parts: one was tested as raw milk, and the other was used for the household boiling experiment. The raw and boiled results come from the same sample, so they are paired measurements, not 240 separate samples. The figure shows where the samples were taken from, and the region codes R01 to R12 were used for regional analysis.

**Fig 1.**
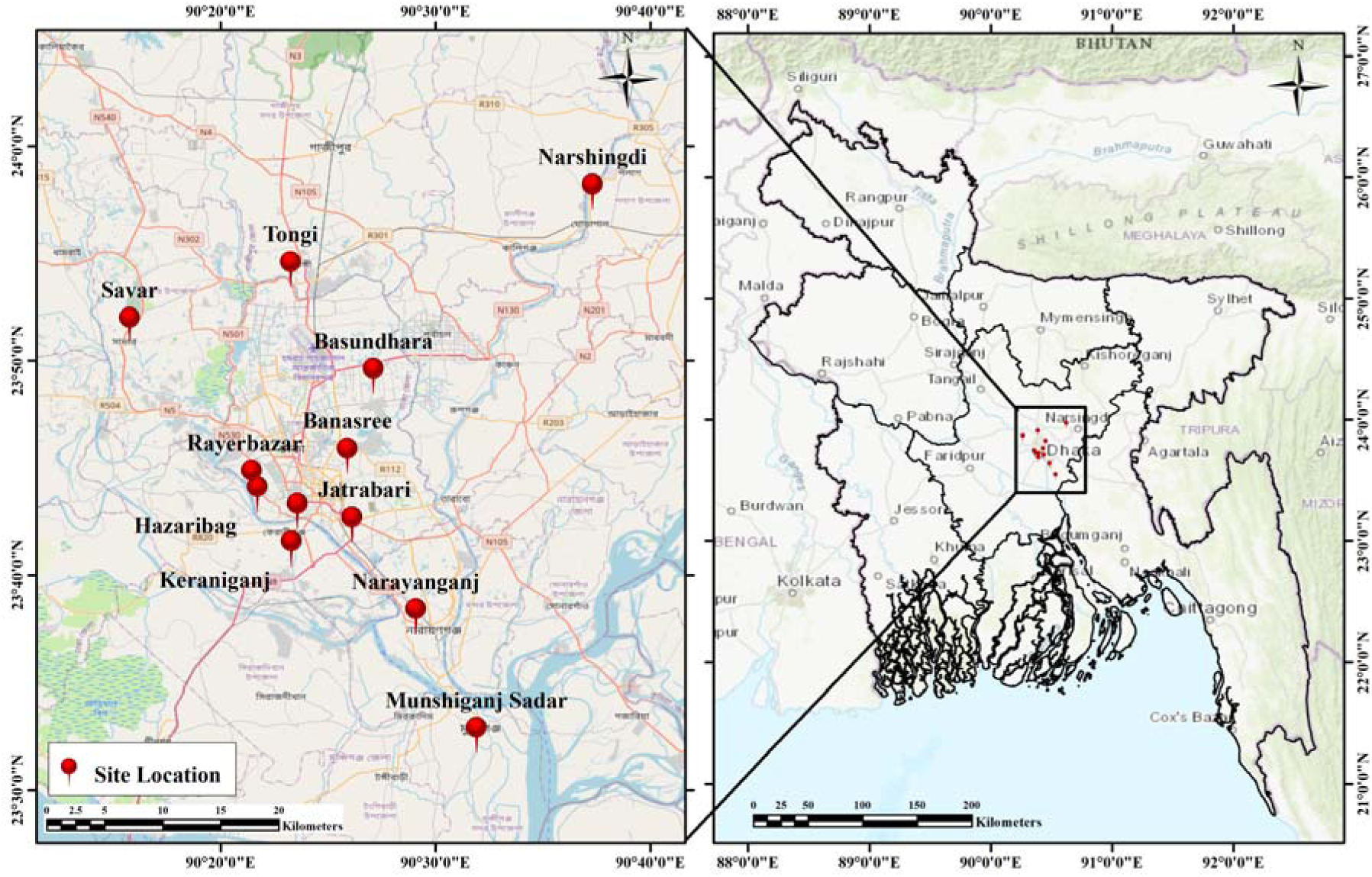
The study area and milk sampling sites are located around Dhaka, Bangladesh. The map was made using ArcMap 10.8 and includes boundary data from GADM. The map also uses an Esri basemap. All the sampling locations are based on data collected during the study.

### 2.2 Ethical considerations and site permission

This study involved checking cow-milk samples from commercial sources. We didn’t have any direct contact with people, handle live animals, do animal experiments, or collect personal information from farmers or vendors. Because of this, ethics review wasn’t needed, and no approval number was given. We got permission from the managers at the milk collection points before taking the samples.

### 2.3 Chemicals, standards, and equipment

Certified reference standards of ciprofloxacin (CIP; CAS 85721-33-1) and enrofloxacin (ENR; CAS 93106-60-6), each with 98% purity, were provided by Sigma-Aldrich/Merck (Darmstadt, Germany) through Incepta Pharmaceutical Ltd. (Dhaka, Bangladesh). The chemicals used included acetonitrile (HPLC grade, Chromasolv®, Sigma-Aldrich/Merck), oxalic acid (Loba Chemie Pvt. Ltd., Mumbai, India), trichloroacetic acid (TCA; research-grade; Research-Lab Fine Chem Industries, Mumbai, India), acetic acid, HPLC-grade water, and deionized water. Chromatography was performed using a Shimadzu Prominence SIL-20 series HPLC system (Kyoto, Japan). The system included an autosampler, dual pumps, a column oven, a degasser, and a UV-vis detector, all controlled with LCsolution software. We separated the compounds on a Luna C18 column (Phenomenex, Torrance, CA, USA; 250 × 4.6 mm, 5 µm). For sample preparation, we used a Hettich Mikro 220R refrigerated centrifuge and a Digisystem VM-2000 vortex mixer.

### 2.4 Stock solutions, blank milk, and matrix-matched calibration

We prepared individual stock solutions at 100 µg/mL, adjusting for 98.0% purity. For each analyte, 10.204 mg was weighed into an amber 100-mL bottle, dissolved first in 0.1 M HCl, and then filled up with HPLC-grade water. These stock solutions were kept away from light, stored at 2-8 °C, and used within a week. A mixed working solution containing 10 µg/mL of each analyte was made with HPLC-grade water, stored in the dark at 2-8 °C, and used within 24 hours.

Blank milk was taken from cows that hadn’t received antibiotics for at least a week or two, and it was checked with HPLC-UV before use. Calibration was done using milk that matched the sample matrix. The blank milk was fortified with known amounts of the analytes at 12.5, 25, 50, 100, and 150 µg/L before extraction. We processed these standards the same way as the actual samples. In the calibration equations, x is the concentration in µg/L of milk-equivalent, and y is the peak area measured. The equations are y = 7148x + 6616 for ENR and y = 6808x + 872 for CIP. The lowest calibration level, 12.5 µg/L milk-equivalent, was used as the minimum level to report.

The purity-corrected standard mass was calculated as:

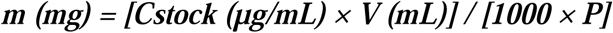

where m is the mass of certified standard, Cstock is the target stock concentration, V is the final volume, and P is the certified purity written as a fraction (0.98).

### 2.5 Milk extraction, concentration calculation, and secondary dilution

For each sample, we gently mixed 2 mL of milk with 8 mL of 5% TCA and vortexed it well. Then, we centrifuged the mixture at 4000 rpm (about 1840 ×g) for 20 minutes using a Hettich Mikro 220R with a 12-place angle rotor. The clear liquid on top was filtered through a 0.45 µm nylon membrane, and 20 µL of this filtrate was injected into the instrument three times as technical repeats. These repeats weren’t counted as separate samples; we only used one reading per aliquot for our overall analysis.

We converted the peak areas into concentrations based on the calibration curve, which accounted for the original milk. Since the calibration solutions were prepared in the same way as the samples, no extra correction was needed.

Out of all the samples, 11 raw milk samples had measurements higher than our calibration curve’s highest point before dilution—3 for CIP and 8 for ENR. None of the boiled samples went over 150 µg/L. For these samples that are outside the normal range, we took 500 microliters of the filtered extract and mixed it with 500 microliters of plain milk matrix, which we prepared using 2 milliliters of milk and 8 milliliters of 5% TCAWe handled this mixture in the same way. We only accepted the diluted extract if its response was between 12.5 and 150 µg/L. Then, we doubled the measured response to find out the final concentration, to account for the dilution.

Dilution integrity was verified at 200 µg/L, with recovery values of 99.1% and a relative standard deviation of 0.9% for ENR, and 99.4% with an RSD of 0.8% for CIP.

The relative centrifugal force was checked from rotor radius and speed using:

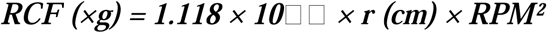

Using r = 10.3 cm and 4000 rpm gives approximately 1840 ×g. Concentrations were calculated from the calibration line as follows:

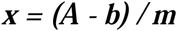

where x is the original-milk matrix-equivalent concentration (µg/L), A is peak area, b is the intercept, and m is the calibration slope.

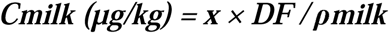

where DF is the secondary dilution factor (1 for undiluted extracts and 2 after the validated secondary dilution) and ρmilk was approximated as 1.0 kg/L.

### 2.6 Household-boiling procedure

A 100-mL portion of each original sample was placed in a 250-mL borosilicate glass beaker and heated without a lid on an electric hot plate. The heat was turned up enough to keep the liquid boiling steadily. We started timing once the boiling was uniform and continued for 15 minutes. During boiling, the milk reached about 100 °C, but we didn’t measure the temperature constantly. After 15 minutes, we removed the beaker from the heat and let the milk cool uncovered until it was about 25 °C. Then, we gently stirred it and took a 2-mL sample for the same TCA extraction as with the raw sample. 2.7 Chromatographic conditions CIP was analyzed using isocratic chromatography with 0.05 M oxalic acid and acetonitrile in a 90:10 ratio. ENR was analyzed with a mixture of 10% acetic acid and acetonitrile in a 90:10 ratio. The flow rate was set to 1.0 mL/min, and each run lasted 20 minutes. UV detection was conducted at 280 nm, with an injection volume of 20 µL. To ensure consistency, reference standards were run regularly to verify retention times, typically around 13.4 minutes for ENR and 16.5 minutes for CIP. System suitability was confirmed when the theoretical plates were at least 2,000, the tailing factor was no more than 2, and replicate injections varied by no more than 2% RSD. Unfortunately, the archived run records did not include the column temperature setting, so we did not verify that value. Because identification depended on matching HPLC-UV response and retention times, the results are preliminary screening estimates rather than confirmed identities by LC-MS/MS [32,33,36].

### 2.8 Analytical method-performance assessment

The method’s performance was evaluated using quality control measures, including matrix-matched calibration, blank-matrix checks, system suitability, precision, recovery, carryover, matrix effect, robustness, dilution integrity, stability, measurement uncertainty, decision-limit information, and batch quality-control summaries. These assessments followed ICH Q2(R2) guidelines and the principles outlined in Commission Implementing Regulation (EU) 2021/808 [53,54]. We conducted recovery tests at 100 µg/L using three separate fortified and extracted blank-milk samples for each analyte. The average recovery was 100.75 ± 0.32% for ENR and 100.28 ± 0.24% for CIP, based on three independent extractions. Intraday precision was 0.38% RSD for ENR and 0.25% for CIP, while interday precision was 0.28% and 0.41%, respectively, across six determinations. Recovery was assessed at a single fortification level rather than at multiple concentrations across the calibration range.

The lowest calibration level used for reporting was 12.5 µg/L in milk equivalent. Results at or above this threshold are considered detected, while lower results are recorded as ND (not detected). This doesn’t mean the compound is completely absent below 12.5 µg/L; it simply indicates that it’s below the detection limit. The estimated limits of detection (LOD) were 0.026 µg/L for ENR and 0.029 µg/L for CIP, with estimated limits of quantification (LOQ) at 0.087 and 0.096 µg/L, respectively. These values are based on regression estimates and are reported separately from the experimentally determined reporting range. Summary validation records are included in S1 File. The following equations were used for the reported validation calculations:

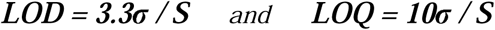

where σ is the response standard deviation used in the original validation calculation and S is the calibration slope. Replicate-level source values used to obtain σ were not available in the archived source package.

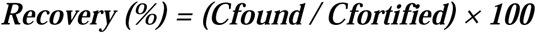

where Cfound is the measured concentration in the fortified blank milk and Cfortified is the nominal fortified concentration.

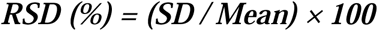

where SD is the standard deviation of the replicate measurements.

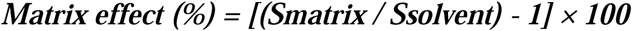

where Smatrix and Ssolvent are the slopes of the matrix-matched and solvent calibration curves, respectively.

**Table 1.** Analytical method-performance summary for enrofloxacin and ciprofloxacin.

| Parameter | ENR | CIP | Interpretation |
| --- | --- | --- | --- |
| Calibration equation | $y = 7148x + 6616$ | $y = 6808x + 872$ | Original-milk matrix-equivalent calibration |
| $R^2$ | 0.999 | 1.000 | Five-point range |
| Evaluated quantitative range | 12.5–150.0 µg/L | 12.5–150.0 µg/L | Lowest level used as operational reporting threshold |
| Calculated LOD | 0.026 µg/L | 0.029 µg/L | Regression-derived estimate; operational reporting threshold = 12.5 µg/L |
| Calculated LOQ | 0.087 µg/L | 0.096 µg/L | Regression-derived estimate; lowest evaluated matrix-matched level = 12.5 µg/L |
| Theoretical plates | 31,628 | 27,412 | Acceptance: $\geq 2000$ |
| Tailing factor | 0.92 | 1.05 | Acceptance: $\leq 2$ |
| Recovery at 100 µg/L | 100.75 ± 0.32% | 100.28 ± 0.24% | n=3 independent extractions |
| Intraday precision | 0.38% RSD | 0.25% RSD | n=6 |
| Interday precision | 0.28% RSD | 0.41% RSD | n=6 |
| Carryover after highest calibrator | <20% of LOQ-level response; no quantification interference | <20% of LOQ-level response; no quantification interference | Summary record |
| Matrix effect | +3.2% (slope ratio 103.2%) | −2.7% (slope ratio 97.3%) | Summary record |
| Robustness | Maximum change 1.4%; RSD 1.3% | Maximum change 1.2%; RSD 1.1% | Summary record |
| Dilution integrity at 200 µg/L | 99.1% recovery; RSD 0.9% | 99.4% recovery; RSD 0.8% | Validated two-fold dilution |
| Expanded uncertainty U (k=2) | 6.8% | 6.2% | Summary record |
| Batch QC near benchmark | 100.6% recovery; RSD 0.7% | 100.2% recovery; RSD 0.6% | Summary record |
| Combined CCα | 105.8 µg/kg | 105.8 µg/kg | Combined ENR+CIP decision characteristic; not the EU MRL |

**Note.** CIP, ciprofloxacin; ENR, enrofloxacin; RSD, relative standard deviation; CCα , decision limit; U, expanded uncertainty. Recovery was evaluated at a single 100 µg/L fortification level.

**Table 2.** Summary stability results under evaluated storage and handling conditions.

| Condition | ENR remaining | CIP remaining |
| --- | --- | --- |
| Fortified milk, room temperature, 6 h | 96.8 ± 1.8% | 97.2 ± 1.6% |
| Fortified milk, 4 °C, 24 h | 97.5 ± 1.5% | 96.9 ± 1.7% |
| Three freeze–thaw cycles | 95.7 ± 2.2% | 96.3 ± 2.0% |
| Processed extract, 24 h | $98.9 \pm 1.1\%$ | $99.2 \pm 0.9\%$ |
| Stock standard, 2–8 °C, 7 d | $99.1 \pm 0.9\%$ | $98.8 \pm 1.0\%$ |
| Working standard, 2–8 °C, 24 h | $99.4 \pm 1.1\%$ | $99.6 \pm 0.8\%$ |

**Fig 2.**
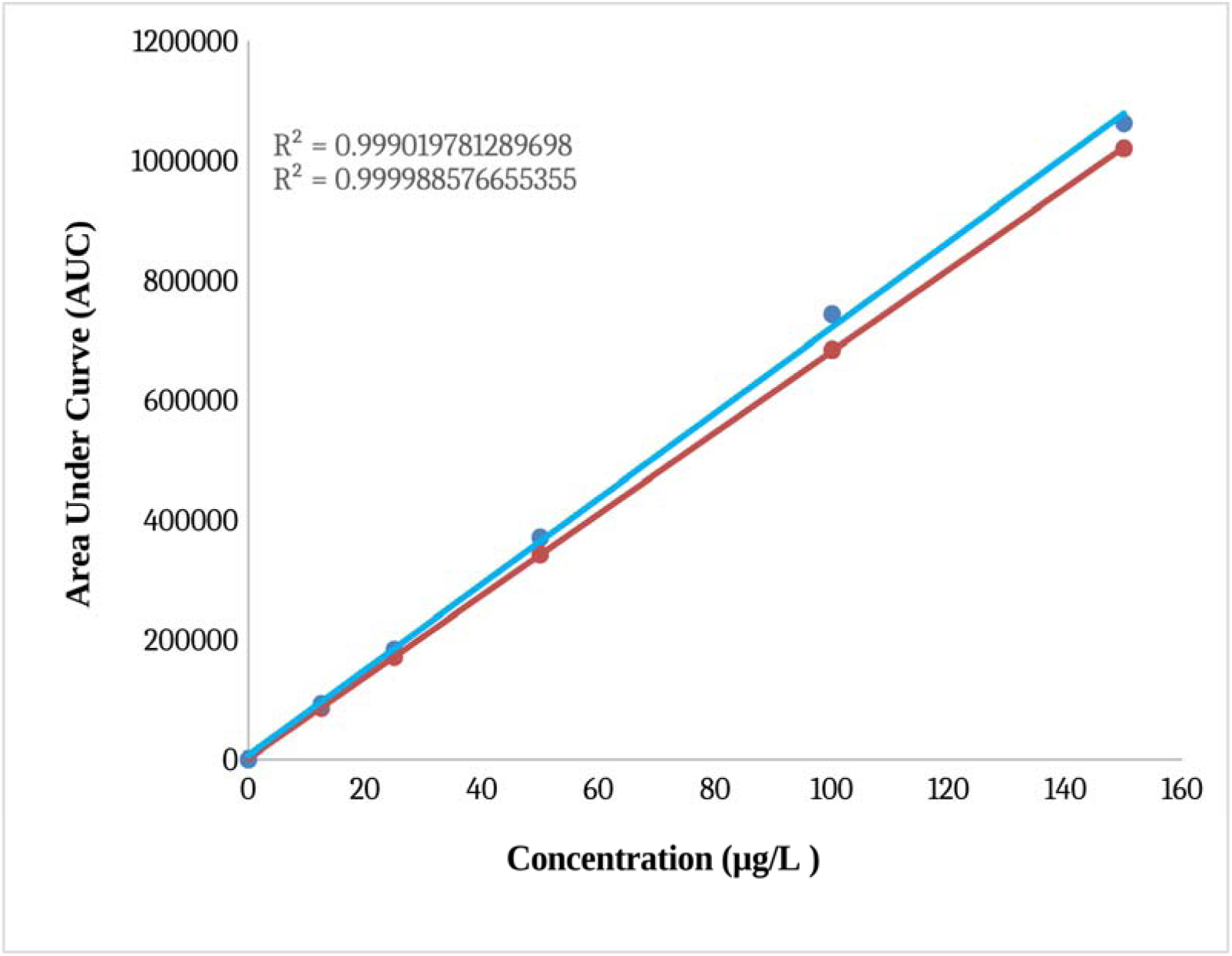
Matrix-matched calibration curves for enrofloxacin and ciprofloxacin. Concentrations on the x-axis represent original-milk matrix-equivalent concentrations. Mean peak-area values underlying the plotted calibration points are provided in S1 File.

**Fig 3.**
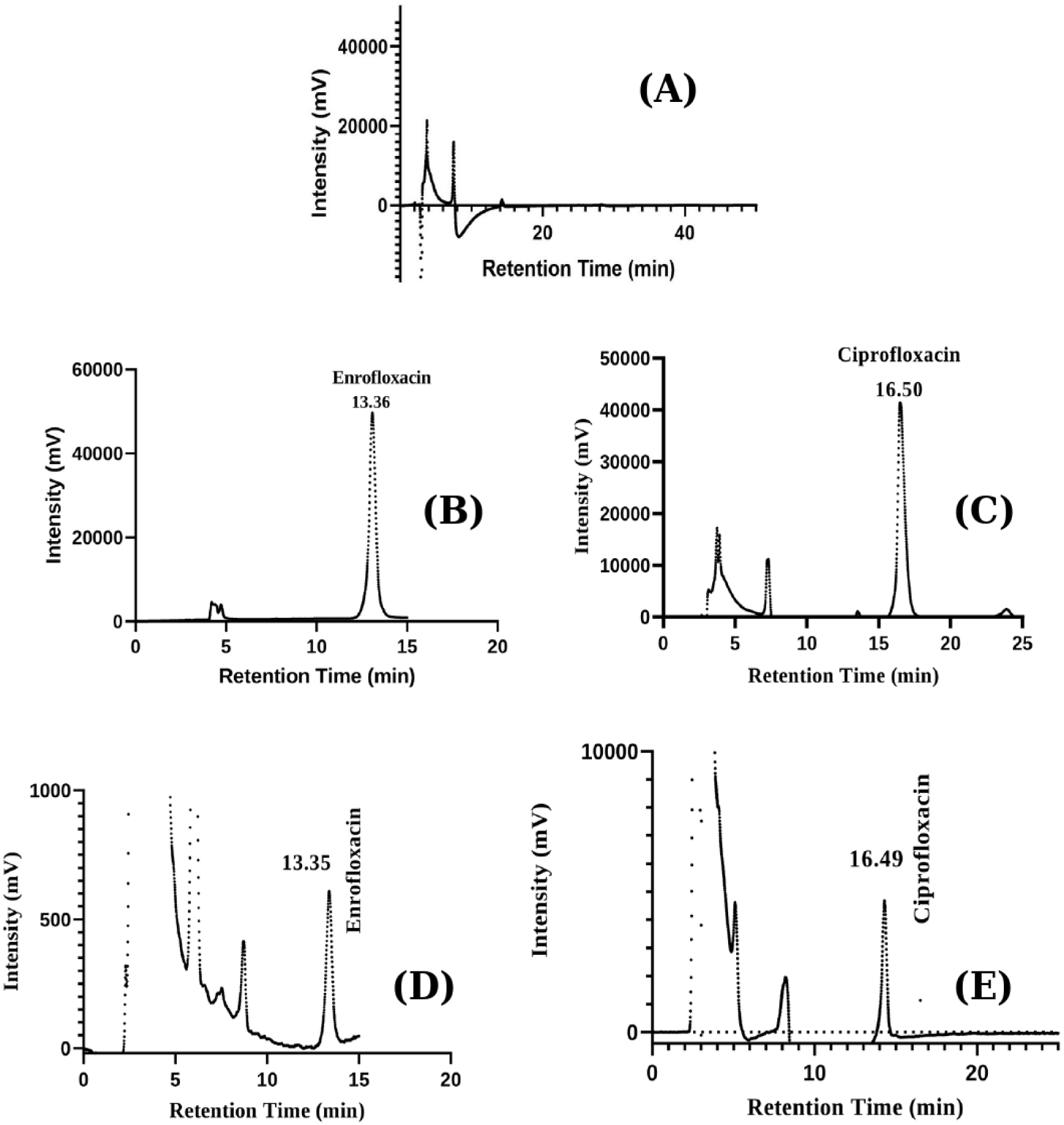
Representative HPLC-UV chromatograms showing (A) blank milk, (B) enrofloxacin reference standard, (C) ciprofloxacin reference standard, (D) a milk extract with an analyte-associated response at the enrofloxacin reference-standard retention time, and (E) a milk extract with an analyte-associated response at the ciprofloxacin reference-standard retention time.

### 2.9 EU MRL benchmark and decision characteristic

The European Union’s limit for the combined amount of ENR and CIP in cow’s milk is 100 µg/kg, which was used as a reference [24]. The test method’s decision limit (CCα) was found to be 105.8 µg/kg.

### 2.10 Screening dietary-exposure assessment

The exposure assessment during screening examined the total measured levels of CIP and ENR. According to the Department of Livestock Services, the average person in the country consumes about 222 mL of milk daily [3]. Since we didn’t have detailed dietary data broken down by age, we used this milk consumption figure as a rough estimate of intake. Since milk weighs about 1 kg per liter, that’s approximately 0.222 kg of milk daily. For adults, we used an average body weight of 60 kg, and for children, about 10 kg, assuming the same intake as a cautious estimate [26].

In this screening, we used the highest acceptable daily intake (ADI) for enrofloxacin set by JECFA, which is 2.0 micrograms per kg of body weight [25]. The hazard index (HI) was calculated by dividing the estimated daily intake (EDI) by this reference value. An HI greater than 1 indicates the exposure estimate is above the reference level. However, it doesn’t necessarily mean there’s a health risk; it just indicates that the estimate exceeds the threshold based on these assumptions [26].

The exposure calculations were:

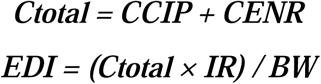

where EDI is estimated daily intake (µg/kg body weight/day), Ctotal is total CIP+ENR concentration (µg/kg), IR is the milk intake proxy (kg/day), and BW is body weight (kg).

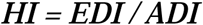

where ADI was 2.0 µg/kg body weight/day for this screening calculation.

### 2.11 Statistical analysis and handling of non-detected results

The original milk sample was the main unit for our analysis. We looked at how the total concentrations of CIP and ENR changed after boiling, focusing on the paired differences. We also measured how often these substances were detected, if they exceeded EU safety limits, how results varied by region, and estimated how much people might be exposed to these residues through their diet. The data analysis was done using IBM SPSS Statistics 26.0, GraphPad Prism 8.4.2.

For detection, anything below 12.5 µg/L of milk equivalent was considered ‘not detected’ (ND). Results at or above this level were considered detected. ND doesn’t mean the substance is completely absent. When summarizing CIP and ENR separately, we ignored ND results and only counted detected samples. For overall residue levels, regional differences, EU benchmarks, and exposure estimates, we treated ND results as zero to include all 120 samples in a lower-bound estimate. We also did a sensitivity test where we replaced each ND with half that level (6.25 µg/kg); detected values stayed the same.

To see if the differences in concentrations were normally distributed, we used the Shapiro-Wilk test. Since the data weren’t normal, we compared the raw and boiled concentrations with Wilcoxon signed-rank tests. To check if samples crossed EU safety limits before and after boiling, we used the McNemar test. We looked at regional differences using Kruskal-Wallis tests, followed by pairwise comparisons with Dunn-Bonferroni adjustment. Results were considered significant if p was less than 0.05. The effect size for the Wilcoxon test was reported as r. Finally, to estimate the confidence interval for the median change in the reproducibility check, we used a bootstrap method with a fixed seed for consistency.

Wilcoxon effect size was calculated as:

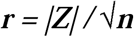

where Z is the Wilcoxon test statistic and n is the number of non-zero paired differences.

### 2.12 Use of artificial intelligence tools

OpenAI ChatGPT was utilized solely for language editing to enhance clarity and readability. It did not contribute to the creation or alteration of research data, statistical results, figures, images, or other primary research materials. The authors carefully reviewed and revised all language suggestions and approved the final version. They bear full responsibility for the accuracy and integrity of the work.

## 3. Results

### 3.1 Analytical performance

The calibration was straight-line across the range of 12.5 to 150 µg/L, with R² values of 0.9990 for ENR and 1.0000 for CIP. Plate counts and tailing factors stayed within the acceptable limits. Both the intraday and interday precision results were very consistent, with RSDs below 1%. At 100 µg/L, the average recovery was about 100.75% for ENR with a standard deviation of 0.32%, and about 100.28% for CIP with 0.24%. More details on method performance and stability are available in Tables 1 and 2. These results support using the method for HPLC-UV screening.

### 3.2 Detection frequency and EU-benchmark status

CIP was found in 113 out of 120 raw samples, which is about 94.2%, close to Wilson’s confidence estimate of 88.4% to 97.1%. The same number of samples, 113 out of 120, also had CIP after boiling. ENR was detected in these same raw and boiled samples. Overall, at least one of these substances was found in 117 out of 120 samples, roughly 97.5%, with a confidence interval of 92.9% to 99.1%. Detection means the amount was at least 12.5 µg/L of milk equivalent; if below this, it doesn’t mean they are completely absent.

In the main analysis, when we considered non-detects as zero, 111 of the raw samples (about 92.5%) and 108 of the boiled samples (around 90%) went over the EU limit of 100 µg/kg. The data showed 108 samples tested positive both before and after boiling, 3 positive only before boiling, none only after boiling, and 9 negative in both. The statistical test gave a p-value of 0.250, meaning boiling didn’t significantly change the chance of exceeding the limit.

**Table 3.** Paired EU MRL benchmark-status table for raw and boiled milk (primary lower-bound analysis).

| Raw >100 µg/kg | Boiled >100 µg/kg | n |
| --- | --- | --- |
| Yes | Yes | 108 |
| Yes | No | 3 |
| No | Yes | 0 |
| No | No | 9 |

**Note.** EU MRL benchmark = 100 µg/kg for the sum of ENR+CIP in bovine milk. Exact two-sided McNemar p = 0.250.

**Fig 4.**
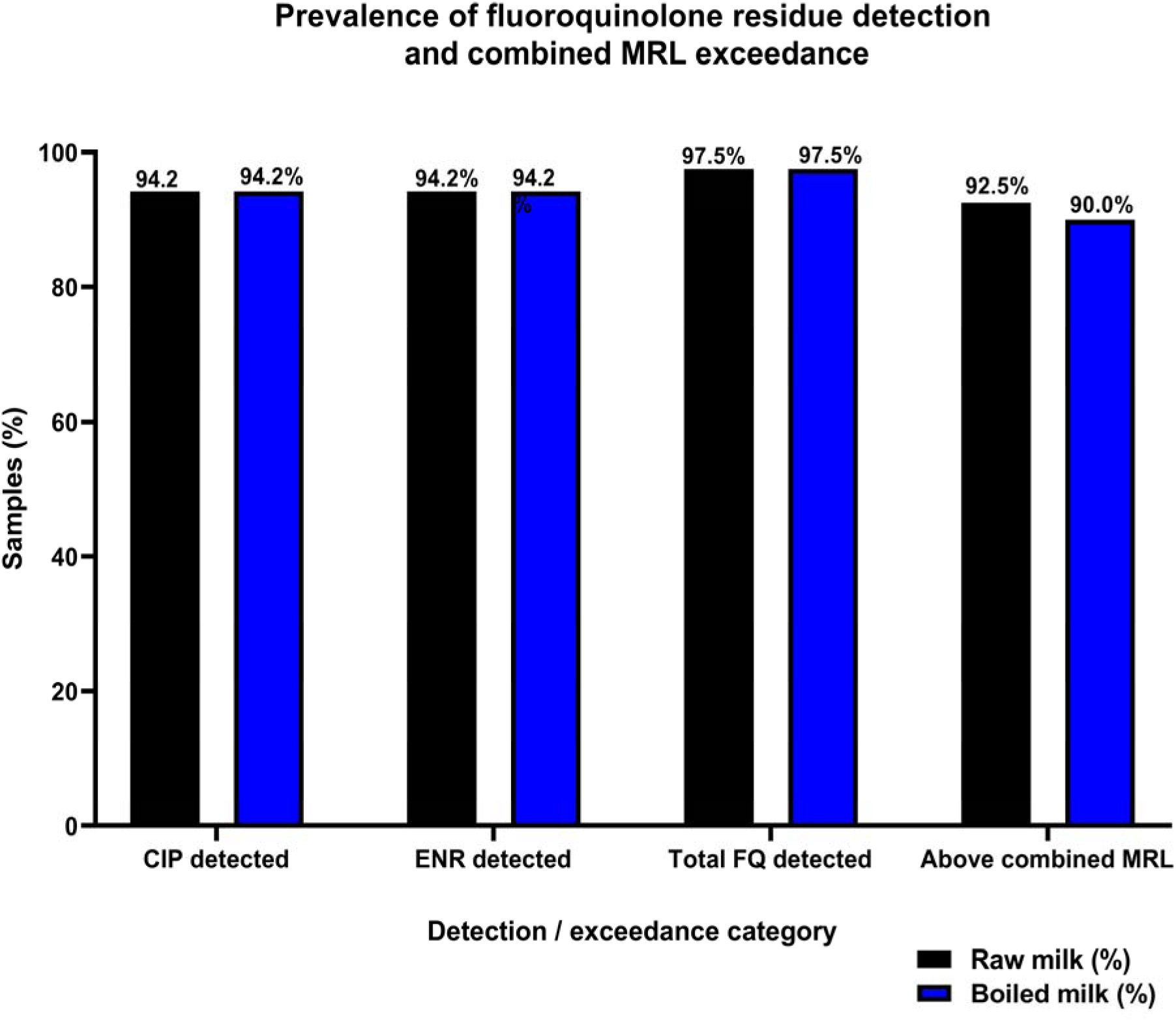
Prevalence of ciprofloxacin and enrofloxacin residue detection and combined EU MRL benchmark exceedance in raw and boiled milk. “Detected” denotes a reportable result at or above 12.5 µg/L milk-equivalent; ND results were below this operational reporting threshold.

### 3.3 Residue concentrations and paired change after boiling

Among the results we found, raw CIP levels ranged from 26.77 to 156.24 µg/kg, with an average of 96.04 ± 28.86, and a median of 98.56. The interquartile range was 40.76. Raw ENR levels ranged from 52.45 to 158.67 µg/kg, averaging 110.62 ± 26.10, with a median of 108.77 and an IQR of 43.89. After boiling, CIP levels ranged from 28.26 to 140.23 µg/kg, with an average of 88.86 ± 26.36, median 95.48, and IQR of 38.25. ENR levels after boiling ranged from 37.32 to 143.45 µg/kg, averaging 101.83 ± 23.74, median 106.32, and IQR 31.31.

Across all 120 original samples in our main analysis, the average total CIP and ENR in raw milk was 194.61 ± 65.72 µg/kg, which dropped to 179.57 ± 59.51 µg/kg after boiling. On average, the difference was approximately −18.19 µg/kg, indicating that levels decreased after boiling. The statistics confirmed this drop was significant, with a Z value of −7.024 and a p-value less than 0.001. When we looked at CIP and ENR separately, both showed lower levels after boiling: CIP dropped by about 10.74 µg/kg, and ENR by about 11.89 µg/kg. These changes were also statistically significant.

**Table 4.** Descriptive concentrations and paired statistical results.

| **Outcome** | **n** | **Mean ± SD**<br>**(µg/kg)** | **Median (IQR)** | **Range**<br>**(µg/kg)** | **Paired result for**<br>**boiled vs raw** |
| --- | --- | --- | --- | --- | --- |
| CIP, raw | 113 | 96.04 ± 28.86 | 98.56 (40.76) | 26.77–156.24 | — |
| CIP, boiled | 113 | 88.86 ± 26.36 | 95.48 (38.25) | 28.26–140.23 | –10.74; Z=–5.104;<br>p<0.001 |
| ENR, raw | 113 | 110.62 ± 26.10 | 108.77 (43.89) | 52.45–158.67 | — |
| ENR, boiled | 113 | 101.83 ± 23.74 | 106.32 (31.31) | 37.32–143.45 | –11.89; Z=–5.940;<br>p<0.001 |
| Total CIP+ENR,<br>raw (ND=0) | 120 | 194.61 ± 65.72 | 204.41 (84.49) | 0–309.91 | — |
| Total CIP+ENR,<br>boiled (ND=0) | 120 | 179.57 ± 59.51 | 195.21 (79.37) | 0–274.42 | –18.19; Z=–7.024;<br>p<0.001 |

Note. Individual-analyte descriptive statistics use detected observations only, where detected denotes a reportable result at or above 12.5 µg/L milk-equivalent. Total-residue descriptive statistics use the primary lower-bound substitution ND=0 and retain all 120 original samples.

**Fig 5.**
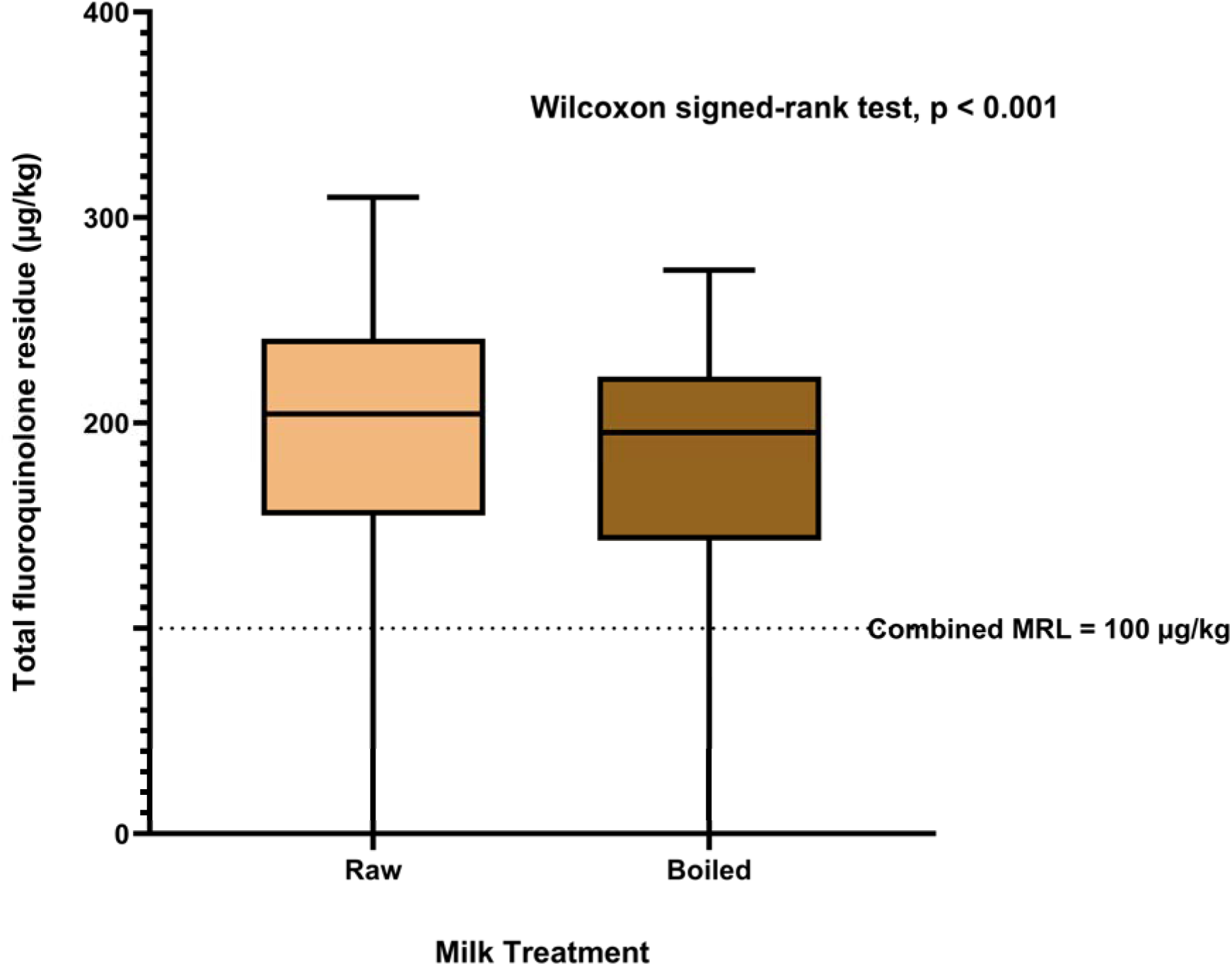
Total fluoroquinolone residues (CIP+ENR) in paired raw and boiled milk samples. The horizontal reference line indicates the EU 100 µg/kg bovine-milk MRL benchmark for the sum of enrofloxacin and ciprofloxacin. ND was assigned zero only for the primary lower-bound analysis.

**Table 5.** Summary of statistical tests and p values used in the main analyses.

| Analysis/comparison | Test/statistic | p value | Interpretation |
| --- | --- | --- | --- |
| Raw vs boiled CIP concentration | Wilcoxon signed-rank test; $Z = -5.104$ | $<0.001$ | Significant decrease after boiling |
| Raw vs boiled ENR concentration | Wilcoxon signed-rank test; $Z = -5.940$ | $<0.001$ | Significant decrease after boiling |
| Raw vs boiled total CIP+ENR concentration | Wilcoxon signed-rank test; $Z = -7.024$ | $<0.001$ | Significant decrease after boiling |
| Raw vs boiled EU-MRL benchmark status | Exact McNemar test | 0.250 | No significant paired change |
| Regional difference in raw CIP | Kruskal-Wallis; $H = 24.905$; $df = 11$ | 0.009 | Significant overall regional variation |
| Regional difference in raw ENR | Kruskal-Wallis; $H = 33.916$; $df = 11$ | $<0.001$ | Significant overall regional variation |
| Regional difference in raw total CIP+ENR | Kruskal-Wallis; H = 30.241; df = 11 | 0.001 | Significant overall regional variation |
| Regional difference in boiled CIP | Kruskal-Wallis; H = 22.234; df = 11 | 0.023 | Significant overall regional variation |
| Regional difference in boiled ENR | Kruskal-Wallis; H = 27.302; df = 11 | 0.004 | Significant overall regional variation |
| Regional difference in boiled total CIP+ENR | Kruskal-Wallis; H = 24.944; df = 11 | 0.009 | Significant overall regional variation |
| Pairwise regional comparisons | Dunn-Bonferroni adjusted post hoc tests | >0.05 after correction | No significant pairwise difference after correction |

Note. CIP, ciprofloxacin; ENR, enrofloxacin; MRL, maximum residue limit; df, degrees of freedom. $p < 0.05$ was considered statistically significant. Sensitivity-analysis results are reported separately in Section 3.5 and Table 7.

### 3.4 Regional variation

Residue levels differed across the 12 study areas for raw CIP, raw ENR, total CIP+ENR, boiled CIP, boiled ENR, and boiled total CIP+ENR. The differences were statistically significant initially, but after adjusting for multiple comparisons, no specific area pairs showed meaningful differences. This suggests that the regional variations are more general rather than highlighting particular areas.

**Table 6.**
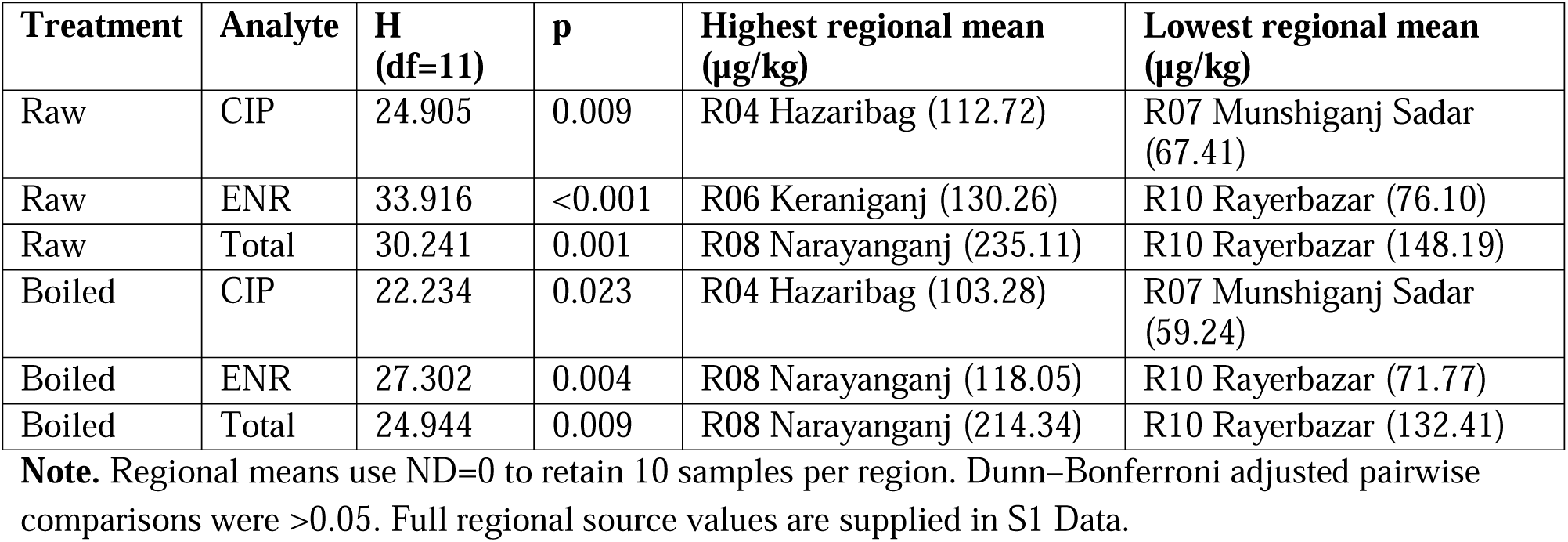
Regional Kruskal-Wallis comparisons and mean-concentration extremes.

| Treatment | Analyte | H (df=11) | p | Highest regional mean (µg/kg) | Lowest regional mean (µg/kg) |
| --- | --- | --- | --- | --- | --- |
| Raw | CIP | 24.905 | 0.009 | R04 Hazaribag (112.72) | R07 Munshiganj Sadar (67.41) |
| Raw | ENR | 33.916 | <0.001 | R06 Keraniganj (130.26) | R10 Rayerbazar (76.10) |
| Raw | Total | 30.241 | 0.001 | R08 Narayanganj (235.11) | R10 Rayerbazar (148.19) |
| Boiled | CIP | 22.234 | 0.023 | R04 Hazaribag (103.28) | R07 Munshiganj Sadar (59.24) |
| Boiled | ENR | 27.302 | 0.004 | R08 Narayanganj (118.05) | R10 Rayerbazar (71.77) |
| Boiled | Total | 24.944 | 0.009 | R08 Narayanganj (214.34) | R10 Rayerbazar (132.41) |

**Note.** Regional means use ND=0 to retain 10 samples per region. Dunn–Bonferroni adjusted pairwise comparisons were >0.05. Full regional source values are supplied in S1 Data.

**Fig 6A.**
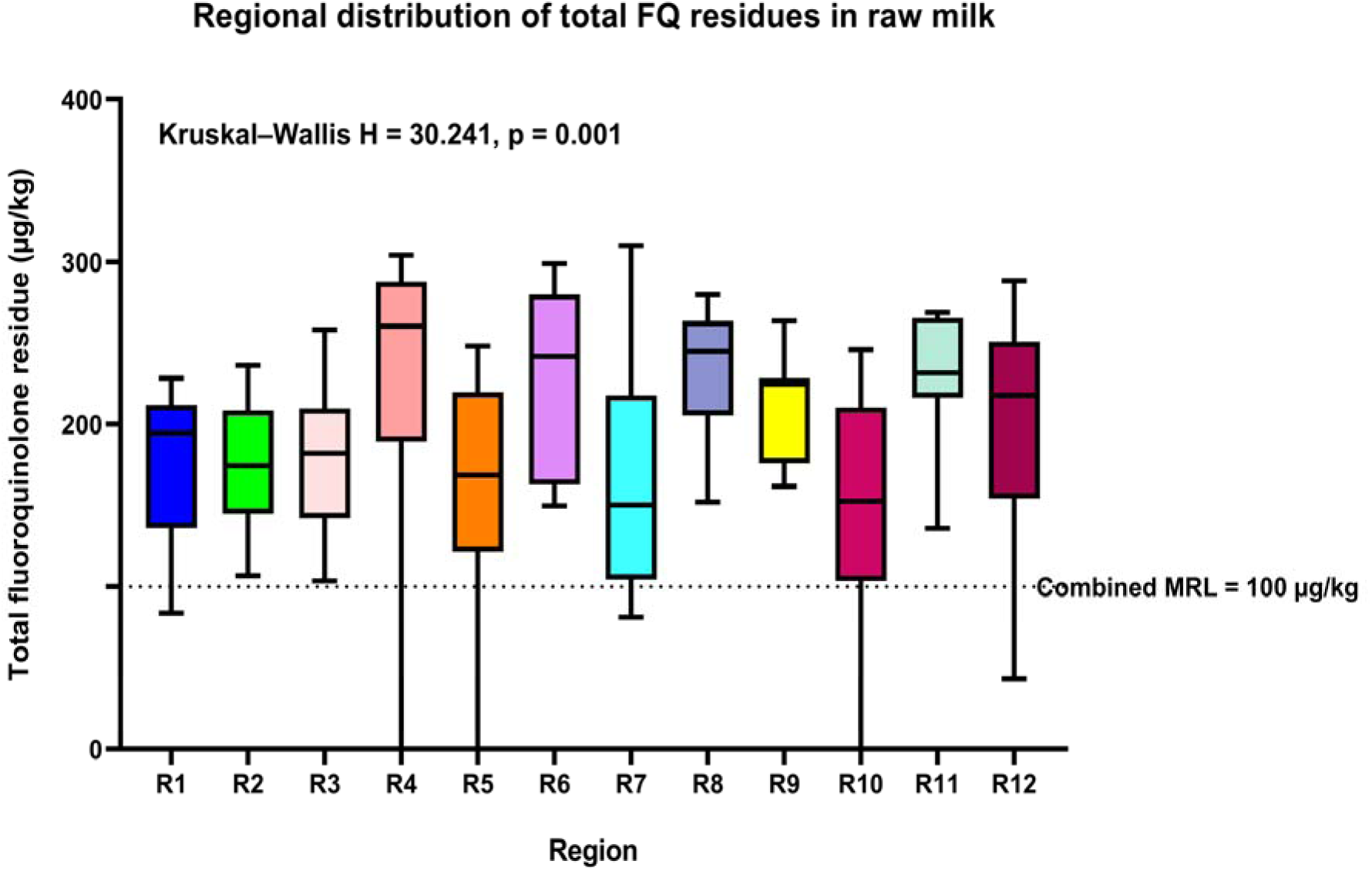
Regional distribution of total fluoroquinolone residues (CIP+ENR) in raw milk samples. Region codes are defined in S1 Data. The horizontal reference line indicates the EU 100 µg/kg benchmark.

**Fig 6B.**
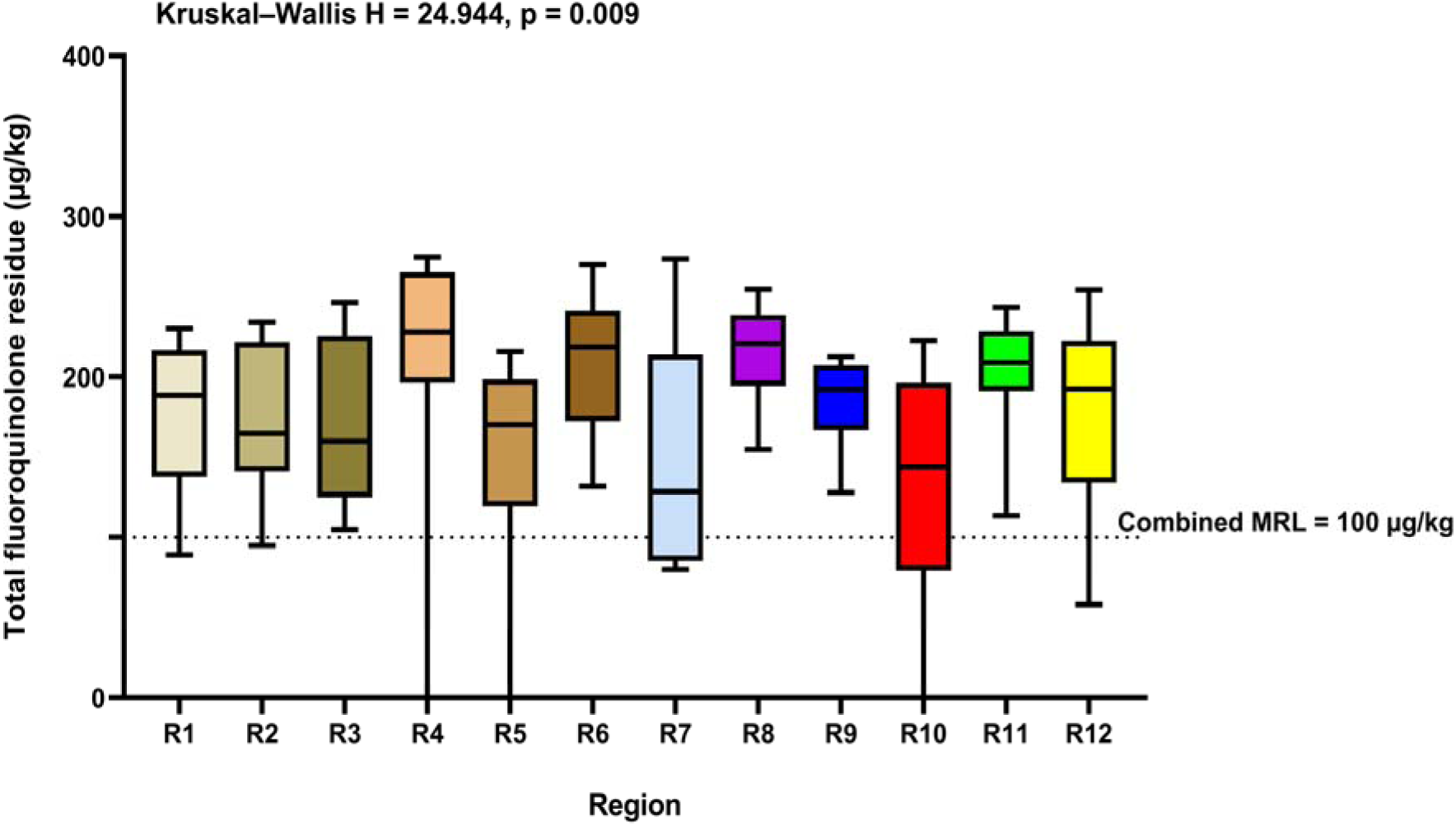
Regional distribution of total fluoroquinolone residues (CIP+ENR) in boiled milk samples. Region codes are defined in S1 Data. The horizontal reference line indicates the EU 100 µg/kg benchmark.

### 3.5 Screening dietary exposure and ND sensitivity analysis

Using the lower estimate of the average total concentration and national milk availability figures as an estimate of intake, we found that an adult’s daily intake was about 0.720 µg per kilogram of body weight for raw milk and 0.664 µg for boiled milk. The hazard indices for screening purposes were 0.360 and 0.332, respectively. For a cautious scenario with a 10-kg child, the intake values increased to 4.318 and 3.984 µg per kilogram, with hazard indices of 2.159 and 1.992.

When we replaced each non-detect result with 6.25 µg/kg, the average concentration changed only slightly—from 194.61 to 195.34 µg/kg in raw milk and from 179.57 to 180.30 µg/kg in boiled milk. The comparison table showed the same results, with 109 yes/yes, 2 yes/no, 0 no/yes, and 9 no/no, with no significant difference (p = 0.500). The regional tests still showed significant differences (raw H = 30.334, p = 0.001; boiled H = 24.998, p = 0.009), and overall, the health risk assessment didn’t change much.

**Table 7.** Screening dietary-exposure estimates under primary and ND sensitivity assumptions.

| ND scenario | Treatment | Population | Mean total (µg/kg) | EDI (µg/kg bw/day) | Screening HI |
| --- | --- | --- | --- | --- | --- |
| Primary ND=0 | Raw | Adult | 194.61 | 0.720 | 0.360 |
| Primary ND=0 | Boiled | Adult | 179.57 | 0.664 | 0.332 |
| Primary ND=0 | Raw | Child sensitivity | 194.61 | 4.318 | 2.159 |
| Primary ND=0 | Boiled | Child sensitivity | 179.57 | 3.984 | 1.992 |
| Sensitivity ND=6.25 | Raw | Adult | 195.34 | 0.722 | 0.361 |
| Sensitivity ND=6.25 | Boiled | Adult | 180.30 | 0.667 | 0.333 |
| Sensitivity ND=6.25 | Raw | Child sensitivity | 195.34 | 4.334 | 2.167 |
| Sensitivity ND=6.25 | Boiled | Child sensitivity | 180.30 | 4.001 | 2.000 |

Note. $EDI = C \times 0.22189 \text{ kg/day} \div \text{body weight}$ . The 0.22189 kg/day value was derived from the national per-capita milk availability figure and was used only as an intake proxy for screening. Adult body weight = 60 kg; conservative child sensitivity body weight = 10 kg. Screening HI = $EDI \div 2.0 \text{ µg/kg bw/day}$ .

## 4. Discussion

### 4.1 Main findings and likely sources of residues

This study found that cow milk from certain dairy regions around Dhaka often contains ciprofloxacin (CIP) and enrofloxacin (ENR), detected through frequent HPLC-UV tests. Out of 120 samples, 113 had CIP and 113 had ENR, with at least one of these antibiotics in 117 samples. On average, the combined amount of CIP and ENR was 194.61 µg/kg in raw milk, dropping slightly to 179.57 µg/kg after boiling. Although the reduction was significant statistically, the number of samples above the EU safety limit stayed roughly the same—92.5% of raw samples and 90% of boiled samples were above 100 µg/kg [24]. This means boiling at home can slightly lower the antibiotic levels, but most samples above the limit before boiling remain above it afterward.

Antibiotic residues in milk usually indicate drug use in dairy animals and suggest treatments may not be carefully managed before milk reaches consumers [5–7,10,13]. Common issues include not following proper withdrawal times, incomplete treatment records, repeated treatments, easy access to veterinary medicines, and limited veterinary oversight [6,7,10,13]. Enrofloxacin is a veterinary drug, and ciprofloxacin can form as a metabolite after ENR treatment [18,22]. Studies show that antibiotic levels tend to decrease over time after treatment, highlighting the importance of timing between drug use and milk collection [23]. This study didn’t include detailed farm records like treatment history, drug doses, or withdrawal periods. So, while these factors likely influence the findings, we can’t confirm them as direct causes.

Another thing to consider is how milk is pooled or combined from different animals or farms before it’s sold [10,13]. This process can mix contaminated milk with clean milk, which can either increase or decrease the residue levels seen in samples [10,13]. This might explain why we see differences across regions, though the study didn’t trace milk back to specific farms. Future studies should look at connecting samples with farm treatment records to better understand where these residues are coming from.

### 4.2 Comparison with previous studies in Bangladesh

The detection rate in this study was higher than in some earlier studies from Bangladesh, but it’s hard to compare directly because of different methods used [12,37,40–43]. These studies varied in sampling styles, types of samples, the number of samples, antibiotics tested, lab procedures, equipment, calibration, and how they defined a positive result. Because of these differences, the number of residues found can really depend on which drugs and testing methods are used [32–34,36,38].

A recent study tested 50 raw milk samples from five farms in Keraniganj and found oxytetracycline in 90% of them, levofloxacin in 66%, ENR in 64%, and CIP in 62% [42]. None of the ENR or CIP samples in that study went over the safety limit [42]. In our study, we detected ENR and CIP by HPLC-UV in 94.2% of samples, many of which exceeded the EU safety standard of 100 µg/kg [24]. These differences could be due to when and where the samples were collected, or to how the results were reported. While earlier work focused only on farm samples and tested 50 samples, we collected 120 samples from different milk collection points in 12 areas. Plus, we looked at the combined levels of ENR and CIP against EU limits rather than each one separately.

Another study by Rinky and colleagues tested 27 samples from Dhaka, including raw, processed, pasteurized, UHT, and flavored milk [43]. They found oxytetracycline in all samples, and 22% had ENR [43]. About 37% of those samples went over the maximum residue limits set in that study [43]. The findings are somewhat different from ours, in which we observed high levels of ENR and CIP. That study examined additional antibiotics and different milk types. Our work focuses specifically on two connected fluoroquinolones in paired raw and boiled samples. Overall, these studies show that antibiotic residues are an issue in Bangladeshi milk across different drugs and product types [37,40–43].

Rahman and colleagues found antibiotic residues in both raw and pasteurized cow milk from Dhaka [41]. This shows that even after processing, milk can still contain antibiotics such as ENR. So, heat-treated milk isn’t necessarily free of these drugs [1,41,47–52]. Another study in Bangladesh looked at how people might be exposed to antibiotics through milk [40,42,43]. It’s not enough to detect residues; we also need to think about the health risks from consuming them. Earlier research from Chittagong found antibiotic residues in milk from both commercial and local farms [37]. Overall, these studies show it’s important to check for residues in Bangladesh regularly. However, because they used different methods, their results shouldn’t be combined into one national estimate.

Anika and team studied how antibiotic levels in milk change over time after cows are treated [23]. They found that residues tend to decrease as the withdrawal period goes on [23]. This highlights how important it is for farmers to follow withdrawal guidelines carefully to keep milk safe [10,13,23]. It also helps explain why testing milk at different times can yield different results—because cows might be at different stages of recovery. Since we didn’t have details on when the cows were treated in our own study, we can only consider this a possible explanation, not a definitive one.

### 4.3 Comparison with studies from other countries

Studies around the world show that milk can contain varying levels of residues, sometimes even comparable to the variations seen across entire countries [32,33,35,36,38,39,44–46]. For instance, Wang and colleagues in Shanghai tested 20 antibiotics in meat, milk, and seafood using advanced mass spectrometry [35]. They found antibiotics in about 11% of milk samples, which was lower than what we found in our study [35]. Their research covered many drug types and used packaged milk, while we focused specifically on ciprofloxacin (CIP) and enrofloxacin (ENR) in milk from our local area. This shows that the two studies looked at different populations and questions.

Another study by Chung and team tested 269 milk samples from cows and goats in Korea [33]. They started with a microbial test that suggested 21 samples might be positive, but when they checked with a more precise method called HPLC, only four samples confirmed the presence of antibiotics like CIP [33]. This highlights how different testing methods can affect what we believe about residue levels. HPLC-UV can give useful estimates, but it’s less accurate than more advanced techniques like LC-MS/MS [32,33,36].

In Kathmandu Valley, Khanal and his team found that HPLC testing detected more positive samples than quick testing kits [38]. They found residues like amoxicillin in 81% of samples, sulfadimethoxine in 41%, penicillin G in 27%, and ampicillin in 12% [38]. This shows that detailed testing can find traces that faster screens might miss, but comparing these numbers across studies can be tricky because different methods are used.

Other studies also looked at milk from different parts of the world. Tasci and colleagues analyzed 130 samples with high-tech LC-MS/MS and found antibiotics or their metabolites in about 55% of them [36]. They didn’t find enrofloxacin, but did find CIP at low levels [36]. Researchers in China tested pasteurized and UHT milk and found no residues above legal limits [45]. In Punjab, India, Moudgil and team found antibiotic residues in raw and commercial milk and even looked into the potential health risks [39]. These studies show that antibiotic residues in milk are a global concern, influenced by local veterinary practices, rules, sample types, and testing methods [32,33,35,36,39,44–46].

The detection rate seems higher here than in most other reports [32,33,35,36,38–46]. But that doesn’t necessarily mean that the dairy industry around Dhaka has more of the issue than other areas. The sites we chose were collection points, not a full-scale national survey, and we used HPLC-UV as a quick screening method without confirming results with more precise LC-MS/MS tests [32,33,36]. So, it’s better to look at overall trends and the need for ongoing monitoring, rather than comparing countries or assuming this is an exact picture of the whole country.

### 4.4 Effect of household boiling and comparison with heat-treatment studies

Boiling significantly reduced the levels of CIP, ENR, and their combined total. The median change was −18.19 µg/kg, and the statistical test showed this result was highly significant (p < 0.001). This suggests that boiling at home can affect the amount of these compounds in milk. However, it doesn’t mean the chemicals were completely destroyed, since the milk was heated uncovered and we didn’t record the initial and final amounts or volumes.

This finding matches earlier studies showing that heat can lower some antibiotic residues but usually doesn’t get rid of them entirely [1,47–52,55,56]. For example, Laszlo and colleagues used a special analysis to look at veterinary antibiotics in boiling raw milk and found some compounds stayed fairly stable [47]. Roca and team specifically looked at quinolones and found that this group can resist heat well [51]. Widiyanti and others also reported that ENR residues can persist even after heating [52]. These studies support our observation that boiling can reduce the measured levels but often leaves behind a significant trace.

How a drug reacts to heat depends on factors like the specific drug, temperature, how long it’s heated, the pH, the milk’s composition, and processing methods [47,49,51,52,55,56]. Fathy and colleagues found that boiling affected residues of oxytetracycline and sulfamethazine in raw milk, showing that antibiotics react differently to heat [56]. Quintanilla’s team studied eight antibiotics in goat milk and cheese, finding some residues can still be present after pasteurization and other processing [55]. For consumers, this means boiling can help kill bacteria and make milk safer, but it shouldn’t be relied on to completely eliminate drug residues [1,47–52,56].

When boiling, evaporation is important. Losing water can make heat-stable compounds seem more concentrated, even if some of them break down [47,49,51,52]. If the measured concentration drops after heating, it could be due to actual breakdown, changes in the food, absorption, redistribution, or a mix of these [47,49,51,52]. Since this study didn’t measure the total weight, the best way to understand what happened is to look at the net change in concentration. Future studies should record the starting and ending weight or volume, see how much residue is left, and use advanced methods like LC-MS/MS to find any breakdown products [36,47].

### 4.5 Regulatory meaning of the EU benchmark

The EU’s MRL of 100 µg/kg for the combined levels of ENR and CIP in cow’s milk was used as a reference point [24]. In the initial analysis, about 92.5% of raw milk samples and 90.0% of boiled samples exceeded this limit. A statistical test showed no significant difference, meaning boiling didn’t much change whether samples were above or below the limit. Most samples that were above 100 µg/kg before boiling stayed above after boiling.

It’s important to understand that this doesn’t mean there’s a legal violation in Bangladesh. The study wasn’t an official inspection, and the EU limit isn’t a legal standard for Bangladeshi milk. Instead, it’s used as an international benchmark. This matters because testing methods and enforcement rules can vary between countries [24,32,54].

Even though many samples exceeded the limit, this info is still useful. It shows there’s a need to manage residue levels in the milk supply better. The best way to do this is to take stronger preventive measures at farms and collection points, such as keeping treatment records, involving vets, following withdrawal periods, and conducting targeted confirmatory tests [6,10,13]. As Berruga and colleagues have emphasized, controlling antibiotic residues starts with proper animal treatment and good milk production practices, not just testing after the milk has entered the market [13].

### 4.6 Dietary exposure and public-health interpretation

The results of dietary exposure should be considered separately from the EU benchmark figures. When using the study’s lower estimate of average total concentration and the country’s per-capita milk availability to guess intake [3], the hazard indices for adults were 0.360 for raw milk and 0.332 for boiled milk — both below 1 given the assumptions. But for a conservative estimate, a 10-kg child showed values above 1. The same national availability figure was used for children and adults, which doesn’t really reflect how much milk Bangladeshi children actually drink [3,26].

This is important to note. The hazard index depends not just on residue levels but also on assumed intake, body weight, and toxicological reference values [25,26]. The figure from the Department of Livestock Services is based on national availability data, not on detailed dietary surveys by age [3]. It’s just a rough estimate for screening. The child scenario shows what happens if the same amount of milk is divided by a much smaller body weight — useful as a safety check, but not a precise measure of risk for children [26].

Overall, the exposure results match previous studies, in which adult hazard quotients or hazard indices remained below 1 even when residues were detected [32,35,39,40,42,43]. Rinky et al. found no immediate risk with their assumptions but noted repeated exposure needs attention [43]. Wang et al.’s Shanghai study used a different approach, including Monte Carlo simulations, and estimated daily antibiotic intake from food at less than 1 microgram per kilogram per day [35]. Their study also showed that contributions from different food groups can vary widely. These comparisons show that exposure estimates depend heavily on the dietary data and model used, not just the measured residue levels [26,35].

MRL exceedance and dietary exposure look at different issues. An MRL mainly relates to approved veterinary drug use, withdrawal periods, and how residues are controlled [24]. A hazard index compares estimated intake to a toxicologically safe reference value [25,26]. So, a milk sample might go over the MRL, but the average adult exposure could still be below the safe limit. These two findings are not contradictory — they address different parts of the food safety picture.

The broader public health concern extends beyond the numerical hazard index. Antibiotic residues in milk can affect sensitive consumers, disrupt dairy starter cultures, and promote antimicrobial resistance [6,14,15,18,57]. Chiesa et al. found that antibiotic residues in raw cow’s milk can affect the lactic acid bacteria essential for cheese-making [15]. Brown et al. reported the presence of both antibiotic residues and resistant bacteria in milk intended for human consumption. However, their study did not establish that one caused the other in specific samples [57]. These insights reinforce the importance of a One Health approach that treats food safety, veterinary drug use, antimicrobial resistance, and environmental impacts as interconnected issues [14,16,17,19,57].

Waste milk and animal waste also play a role. If not properly managed, antibiotic-laden waste can contaminate wastewater, soil, and surface water [16,17,19]. Studies have shown that treatments such as ozonation can break down antibiotics in water and waste milk, underscoring the need for environmental residue management [19]. While such treatments are not a substitute for careful veterinary practices, they highlight the importance of addressing residues after they leave the animal and before the milk reaches consumers.

### 4.7 Regional variation

The Kruskal-Wallis tests showed that there are differences across the 12 study areas in the levels of raw and boiled CIP, ENR, and total CIP+ENR. But after adjusting for multiple comparisons with the Dunn-Bonferroni correction, no specific pairs of regions were found to be significantly different. This means there’s some variability, but we can’t confidently say that any one area is truly different from another.

Several factors might explain regional differences, such as the types of suppliers, disease pressures, access to veterinary care, drug choices, treatment routines, withdrawal practices, or pooling methods at collection points [7,10,12,13]. We didn’t measure these factors directly, so this analysis is just a starting point. With only 10 samples from each area, the main goal is to guide future sampling efforts rather than definitively classifying areas as high- or low-risk.

In future studies, repeated sampling and clearer information on the number and types of milk sources will be helpful. Connecting residue findings with farm practices can provide better insights. Including seasonal sampling is also useful, since disease patterns and antibiotic use can change throughout the year [50]. For national estimates, using a proper, representative sampling method will lead to more accurate results.

### 4.8 Analytical context, strengths, and limitations

The methods used in this study have some real benefits. The calibration was customized for the milk samples and done using original milk. Both the test samples and the calibration standards went through the same extraction process. When responses were above the detection limit, reanalysis was performed after diluting the sample twofold, and recovery was checked with three separate blank milk samples fortified at 100 µg/L. The study treated technical repeats and the original milk samples as main units for analysis and also included a sensitivity check for non-detected values.

Many different methods have been used to test for antibiotic residues in milk, from simple microbiological tests to advanced techniques like HPLC with fluorescence, diode-array, and mass spectrometry detection [20,27–34,36]. For example, Samanidou and colleagues used an HPLC-DAD method with fabric-phase sorptive extraction to find penicillins in milk [29]. Xu and others used HPLC-UV after microextraction to detect tetracyclines [30]. Mohebi and his team also developed an HPLC method after liquid-liquid microextraction for various antibiotics [31]. These studies show that HPLC methods can be useful for routine testing, but their accuracy depends on how well the extraction, calibration, and selectivity are done, as well as the detection limits needed [27–31].

One important point is that our study identified antibiotics based only on HPLC-UV response and retention time, without confirming with more specific methods like LC-MS/MS [32,33,36]. This is especially important because we tested frequently. A confirmatory test that is more selective would help reduce uncertainty caused by substances in the milk that might look similar [32,33,36]. So, our results should be seen as screening estimates rather than confirmed counts of antibiotics in the milk. This cautious view matches what others, like Chung and colleagues, have said about screening versus confirmation [33].

There are also other limits to our study. We collected samples only from certain points around Dhaka, so the findings might not reflect the whole country. Recovery tests were done at just one level, not across different concentration ranges. We didn’t have details about the grade or brand of acetic acid or the conditions of the columns used. We also didn’t measure the mass or volume before and after boiling. We only tested two antibiotics, CIP and ENR, so other antibiotics might have been present but weren’t checked. Plus, the exposure estimates used fixed body weights and national milk availability as a rough guide for intake, instead of real age-based consumption data [3,26].

Despite these limits, the way we paired raw and boiled samples from the same milk helped us minimize differences and made our main comparisons more meaningful. This allowed us to analyze the effects of household boiling more accurately. The sensitivity check, which used half the lowest detection level for non-detected cases, showed that our main conclusions didn’t depend on just assuming the lowest possible value for undetected samples.

### 4.9 Implications for surveillance and future research

The findings show how important it is to stop drug residues from reaching consumers in the first place. Key steps include better veterinary oversight, clear treatment records, farmer education, following withdrawal periods, screening milk at collection points, and targeted confirmatory testing for regulation [6,10,13]. Boiling household milk can still be recommended for microbiological safety when needed, but it doesn’t reliably remove fluoroquinolone residues [1,47–52].

A practical way to monitor this could be in two steps. First, low-cost screening can spot samples or supply points that need more testing [33,34]. Then, more accurate testing with LC-MS/MS can be done when important decisions or regulations are involved [32,36,54]. This approach builds on existing studies that combine quick screening with more precise lab tests [32–34,36]. It helps labs use their resources wisely and trust their results more.

Future research should go beyond Dhaka, include repeated seasonal sampling [50], and use random sampling if we want to understand the whole country. Recording treatment details—drug names, doses, withdrawal times, and farm sources—should be a priority [10,13]. Using multi-residue LC-MS/MS can help study CIP and ENR along with other veterinary drugs like antibiotics and sulfonamides [32,36,45]. Boiling tests should check residue levels before and after heating to tell if they’re lost through evaporation or are truly gone [47,49,51,52]. Better dietary surveys are also needed to accurately measure exposure for adults and children based on what people actually eat, not just what’s available [26].

This study provides new data on household boiling and fluoroquinolone residues in Bangladeshi milk. Its main point isn’t that boiling completely destroys residues, but that it slightly reduces CIP+ENR levels with little effect on safety standards. So, to control residues, the focus should be on better veterinary practices and the dairy supply chain before the milk gets to homes [10,13].

## 5. Conclusion

This study looked at 120 cow-milk samples from different dairy areas around Dhaka. Each sample was divided into two parts: one raw and one boiled at home. Testing with HPLC-UV showed that many samples had detectable levels of CIP and ENR. The average amount of these antibiotics dropped from 194.61 µg/kg in raw milk to 179.57 µg/kg after boiling. This decrease was statistically significant, but the difference wasn’t large enough to change how we see the safety levels. Most samples—92.5% raw and 90% boiled—still went over the EU limit of 100 µg/kg. The test showed no significant difference between raw and boiled samples.

These results suggest boiling the milk at home can slightly reduce antibiotic levels but doesn’t fully remove the concerns about residue. Previous research also shows that antibiotics like quinolones can stay fairly stable even after heating [47,51,52]. Since the milk was boiled uncovered and we didn’t measure its weight before and after boiling, the numbers mainly show concentration changes, not chemical breakdown. Boiling helps kill germs but shouldn’t be seen as a way to get rid of antibiotic residues that come from how the milk was produced [1,10,13,47–52].

This study explains why we should look at regulatory results and exposure risks separately [24–26]. The fact that many samples exceeded the EU limit shows there are issues with residue control in the supply chain. The hazard index for adults stayed below 1 in our assumptions, but for a 10-kg child, it went over 1 in a conservative scenario—though that used the same milk consumption as adults, so it’s more of a safety check than a real estimate for kids. To better understand the risks, we need data on how much children actually drink and more detailed info on combined exposure to ENR and CIP.

When we compare with earlier studies from Bangladesh and other countries, we see that antibiotic residue levels can vary a lot. Past Bangladeshi studies found antibiotics like ENR, CIP, tetracyclines, and others in different types of milk—raw, processed, pasteurized, UHT, flavored—but the rates and how often they exceeded safety limits differ from what we found here [12,37,40–43]. International research shows similar differences, influenced by veterinary practices, milk sources, testing methods, and reporting standards [32,33,35,36,38,39,44–46]. So, the high levels we found should be seen as a regional snapshot using HPLC-UV, not as proof of a national problem.

This study highlights the need for better measures to control and check for residues in the dairy supply. Better veterinary oversight, clear treatment records, following withdrawal periods, screening at collection points, and confirmatory tests with more advanced tools like LC-MS/MS are essential—more than just boiling the milk at home [10,13,32,34,36]. Future studies should include more diverse samples, seasonal checks, detailed treatment histories, multi-residue testing, measurements of milk before and after boiling, and age-specific consumption data. Within these limits, our findings show that boiling can slightly lower CIP and ENR levels but doesn’t reliably bring most high-residue samples below safety limits.

## Data availability

All the data collected at the sample level that support this study’s findings are included in the Supporting Information. S1 Data offers the full dataset for the 120 original milk samples, detailing paired raw and boiled CIP and ENR results, sampling locations, and variables used in statistical, regional, EU-benchmark, sensitivity, and dietary-exposure analyses. The same dataset is also available in CSV format for easy use with standard statistical software.

## Author contributions

**Conceptualization:** Asma Rahman, Munir Ahmed, Md. Abdullahil Asif, Farhana Rinky, Md. Golam Mostafa, Mohammad Nazrul Islam Bhuiyan, Sadia Afrin, Taihan Ahmed. **Data curation:** Asma Rahman, Md. Abdullahil Asif, Farhana Rinky. Formal analysis: Asma Rahman, Md. Abdullahil Asif, Farhana Rinky. **Investigation:** Munir Ahmed, Asma Rahman. **Methodology:** Asma Rahman, Md. Abdullahil Asif, Farhana Rinky, Sadia Afrin, Taihan Ahmed. **Project administration:** Munir Ahmed, Md. Golam Mostafa, Mohammad Nazrul Islam Bhuiyan, Asma Rahman. **Resources:** Asma Rahman, Munir Ahmed. **Software:** Md. Abdullahil Asif, Farhana Rinky, Asma Rahman. **Supervision:** Asma Rahman. **Validation:** Asma Rahman**. Visualization:** Asma Rahman, Munir Ahmed, Md. Abdullahil Asif, Farhana Rinky. **Writing – original draft:** Munir Ahmed, Asma Rahman, Md. Abdullahil Asif, Farhana Rinky. **Writing – review & editing:** Asma Rahman, Munir Ahmed, Md. Golam Mostafa, Mohammad Nazrul Islam Bhuiyan.

## Acknowledgments

The authors thank the staff and laboratory assistants at the Food, Nutrition and Agriculture Research Laboratory, Centre for Advanced Research in Sciences (CARS), University of Dhaka, for their support during sample analysis.

## Supporting information captions

S1 Data. Complete sample-level dataset and analysis workbook, including data dictionary, dilution factors, statistical inputs, regional tables, figure-source values, ND sensitivity analysis, McNemar audit, Wilcoxon audit, and worked concentration calculations (XLSX).

S2 Data. Machine-readable sample-level dataset used by the reproducibility script (CSV).

S1 File. Analytical method-performance summary, stability summary, calibration-figure source means, secondary-dilution records, and method metadata (XLSX). This file contains summary-level records and does not reconstruct missing raw instrument or replicate-level validation data.

